# Survival of hospitalized COVID-19 patients in Northern Italy: a population-based cohort study by the ITA-COVID19 Network

**DOI:** 10.1101/2020.05.15.20103119

**Authors:** P. Giorgi Rossi, E. Ferroni, S. Spila Alegiani, G. Pitter, O. Leoni, D. Cereda, M. Marino, M. Pellizzari, J. Sultana, G. Trifirò, M. Massari, the ITA-COVID19 working group

## Abstract

**Background:** COVID-19 case fatality rate in hospitalized patients varies across countries and studies, but reliable estimates specific for age, sex, and comorbidities are needed to design trials for COVID-19 interventions. Aim of this study is to provide population-based survival curves of hospitalized COVID-19 patients.

**Methods:** A cohort study was conducted in Lombardy, Veneto, and Reggio Emilia using COVID-19 registries linked to hospital discharge databases containing patient clinical histories. All patients with positive SARS-CoV-2 RT-PCR test on oral/nasopharyngeal swabs hospitalized from 21^st^ February to 21^st^ April 2020 were identified. Kaplan Meier survival estimates were calculated at 14 and 30 days for death in any setting, stratifying by age, sex and Charlson Index.

**Findings:** Overall, 42,926 hospitalized COVID-19 patients were identified. Patients’ median age was 69 years (IQR: 57-79), 62·6% were males, 69·4% had a Charlson Index of 0. In total, 11,205 (26·1%) patients died over a median follow-up of 24 days (IQR: 10-35). Survival curves showed that 22·0% of patients died within 14 days and 27·6% within 30 days of hospitalization. Survival was higher in younger patients and in females. Younger patients with comorbidities had a lower survival than older ones with comorbidities.

**Interpretation:** Over 27% of hospitalized COVID-19 patients died within one month in three areas of Northern Italy that were heavily affected by SARS-CoV-2 infection. Such a high fatality rate suggests that trials should focus on survival and have follow-up of at least one month.

**Funding:** The study did not receive any external funding.

**Research in context:** Evidence before this study

Two recent systematic reviews with meta-analyses report case fatality rates of three to four percent in COVID-19 patients. Most studies on hospitalized cohorts report only slightly higher figures. These figures do not correspond to those derived from routinely collected clinical data in most European countries, reporting a 10% case fatality rate which has been increasing over time since the epidemic started.

Robust and precise survival estimates of hospitalized COVID-19 patients which take into account prognostic factors such as age, sex and burden of comorbidities are needed to design appropriate phase II and phase III clinical studies of drugs targeting COVID-19.

**Added value of this study:** In this study we present the first survival estimates by age, sex and Charlson index for a large population-based cohort of Italian hospitalized COVID-19 patients.

**Implications of all the available evidence:** Over 27% of COVID-19 patients died within one month from hospital admission. Such a high fatality rate suggests that studies should prioritize mortality as primary outcome. Furthermore, we found that the fatality rate reaches a plateau 30 days after hospitalization, suggesting that studies should have at least one month of follow up to observe deaths; shorter follow-up could lead to overestimation of treatment benefits.

## Introduction

The novel SARS-CoV-2 has caused a pandemic in early 2020. The virus has shown a high reproduction number and has spread rapidly on a global scale,^1-2^ with more than three million cases of coronavirus disease (COVID-19) being diagnosed worldwide.^3^ Italy was one of the first countries to face the epidemic outside of China.^4-5^ As of the end of April 2020, Italy is second only to the USA as the country with the highest number of COVID-19-related deaths, with a total of around 30,000 deaths and a case fatality rate close to 13·6%.^3^

The spectrum of COVID-19 disease ranges from asymptomatic to severe mixed interstitial-alveolar pneumonia, that may lead to severe acute respiratory distress syndrome and death.^6^ While several important epidemiological findings have emerged about the pandemic, the COVID-19 case-fatality rate (CFR) remains unknown. Reports from different countries show an enormous heterogeneity, ranging from less than 1% to approximately 12% CFR.^3,7^ The accuracy of this assessment, however, is limited by disease ascertainment challenges, bias towards symptomatic and very sick patients, and variability in testing accuracy.^8^ An important source of this heterogeneity is very likely the difference in case detection and reporting (denominator) and also how COVID-19-related deaths are defined (numerator). Furthermore, the length of observation time is key to accurately measure CFR.^9-11^ Where widespread screening was performed in the general population (e.g., in South Korea), the overall CFR is obviously lower, because the denominator includes many mild or asymptomatic cases. However, in countries where mainly people presenting to the emergency department are being screened (e.g., in the Italian region of Lombardy, during the strongest wave of the epidemic), CFRs are higher, because the denominator will include predominantly severe cases.^12^

Two recent systematic reviews of trials and observational studies, including studies mainly from China, found a CFR pooled estimate of 3-4%;^13,14^ however, most reports from European countries show CFRs ranging from 10% to 20% when a cohort approach with adequate follow up was conducted.^9,15^ Differences in CFR are also reported in studies including only hospitalized COVID-19 patients.^16-20^ Reliable population-based estimates of the CFR for hospitalized patients are essential for providing public health standards to health care providers that have to monitor the impact of the epidemic and disease management. CFRs can also be useful to provide a reference standard to accurately design trials of interventions targeting COVID-19 with adequate statistical power. Having a reliable expected number of deaths in a cohort of hospitalized COVID-19 patients with known age and comorbidity burden is critical to design adequately-powered experimental studies and, in particular, to design phase two, single-arm studies, that are currently very frequently employed in ongoing COVID-19 intervention trials. In particular, 729 interventional studies on COVID-19 patients are registered in clinicaltrials.gov as of 30^th^ April 2020 (supplementary figure 1 and table 1). Of these, 37·9% are phase II (including phase I/II and II/III) trials and one third of those studies are aimed at exploring survival as outcome (supplementary figure 2).

Given the rapid spread of the pandemic and the absence of any effective therapy, it is likely that new therapies, if also proven to be promising in phase II non-randomized studies, will be directly transferred to clinical practice without more rigorous testing of their efficacy in phase III studies. The aim of this study is therefore to provide population-based survival curves of hospitalized COVID-19 patients that can be used to monitor the impact of epidemic and disease management, to design phase II trials and to correctly calculate the required sample size of phase III studies.

## Methods

### Setting

The study included patients living in the Lombardy region, first and the hardest hit Italian region (10 million inhabitants), the Veneto region, where the second epidemic wave broke out (4·9 million inhabitants,) and in Reggio Emilia Local Health Unit, which is part of the Emilia-Romagna region, ranking second for the spread of the epidemic during the study period (0·5 million inhabitants). Together, the areas included in the study represent 25% of Italian population.

In Italy the National Health System (NHS) provides all testing activities and acute care free of charge for all residents. All RT-polymerase chain reaction (PCR) SARS-CoV-2 tests performed in Italy must be recorded in a dedicated COVID-19 surveillance registry. Regional health systems are organized in different ways and testing strategies differ based on local protocols and logistic constraints during the study period: in Veneto contact tracing and tests among asymptomatic contacts as well as patients with mild symptoms was carried out since the epidemic began and continued over time. Lombardy had the fastest progression of the epidemic and testing outside the hospital setting quickly became soon untenable, while Reggio Emilia faced an intermediate situation. In all the study catchment areas, patients with symptoms potentially suggesting COVID-19, who were admitted to the Emergency Room (ER) or were admitted to hospital, were always tested.

### Study population

Data were retrieved from the COVID-19 surveillance registry coordinated by the National Institute of Health and implemented in each catchment area. This registry collects information about symptoms, diagnoses, hospitalizations, intensive care unit (ICU) admission, death, and recovery concerning patients testing positive for SARS-CoV-2 RNA by RT-PCR on nasopharyngeal or throat swab samples. Information was collected by the public health departments of the local health authorities from different sources, including molecular laboratories, hospitals, and death certificates and reported to the regional surveillance system within 48 hours (figure 1).

**Figure 1.**
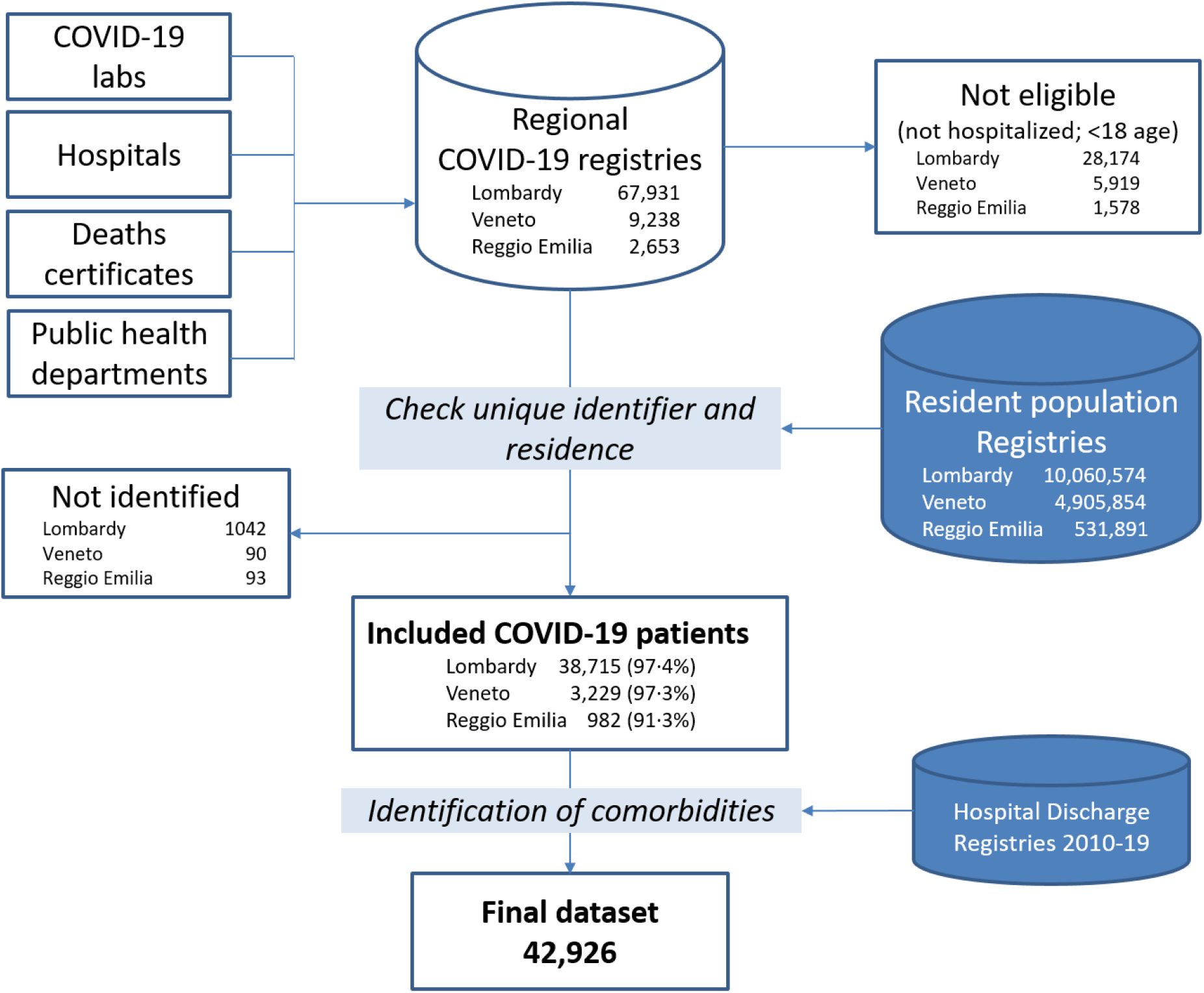
Flowchart of data sources used in the present study. COVID-19 registries at the regional level collect, for all SARS-CoV-2 RT-PCR positive patients, data on date of diagnosis, symptoms, hospitalizations, death, and recovery. These registries were linked to the resident population registries. We report the percentage of patients successfully linked; in some cases, deterministic linkage could not be carried out, likely due to patients not residing in the region or to inaccurate individual data reporting in the COVID-19 registry. Patients confirmed to be residents in the region of interest were found using data from prior contact with the NHS. This data was used to identify comorbidities.

From the COVID-19 registry all patients aged ≥18 years with SARS-CoV-2 infection who were hospitalized from 21^st^ February (date of the first Italian COVID-19 patient hospitalization) to 21^st^ April 2020 were identified in Lombardy. Patients were identified from 21^st^ February to 31^st^ March 2020 in the other two catchment areas. The admission date was considered the index date (ID). Patients were followed up until death or the end of available data, whichever came first. Hospital discharge databases were linked to the COVID-19 registry to identify hospital admissions in the 10 years preceding COVID-19 hospitalizations in order to calculate the Charlson index for each patient.^21^

An R-based tool for distributed analyses developed by the National Institute of Health (The ShinISS) was used by each study center to locally analyze COVID-19 patient data using a common data model, sharing only an anonymized dataset for central analysis, in compliance with EU-GDPR regulations.

### Outcome measure

The end-point was death occurring during follow-up in an inpatient or outpatient setting for any cause, as reported in COVID-19 registry. The outcome measure was time to event from COVID-19 hospitalization to death or the end of available follow-up, whichever came first.

### Covariates of interest

Besides sex, age and catchment area, Charlson index was calculated in order to take into account the impact of the comorbidity burden.

### Statistical analysis

Univariate survival measures, with corresponding 95% confidence intervals (95% CI), at a fixed follow-up time (14 and 30 days) were computed with the Kaplan Meier estimator, for each covariate: age (<50, 50–59, 60–69, 70–79, 80–89, and ≥90 years), sex (males and females) and Charlson index (0, 1–2, and ≥3). Furthermore, we also present data for sex and Charlson index, stratified by age category. A Cox proportional hazard model was used to show the effect of sex, adjusted for age and comorbidities, with an interaction term between age and Charlson index.

### Ethics

The study protocol was approved by Ethics Committee of the Italian National Institute of Health on 18^th^ March 2020 and subsequently by local Institutional Review Board or Ethics Committee.

### Data sharing statement

Data will be updated periodically. Aggregate data are available upon submission of a request which describes the research objectives and a protocol for analyses to the local competent authorities: for Lombardy, queries should be submitted to a specific COVID-19 Committee, identified by the Directorate General for Health (under DDG n. 3019/2020); for Veneto to the Health Direction of the Azienda Zero; for Reggio Emilia to the Comitato Etico Area Vasta Emilia Nord, Reggio Emilia office. Data will be available at least for seven years from the approval of the study.

## Results

The three regional COVID-19 registries captured data from 79,882 patients with at least one positive PCR result over the study period; 35,671 patients were not hospitalized or were below 18 years old. Finally, 1,243 patients were excluded because their data could not be deterministically linked to the resident population registries, resulting in a 97·2% success of record linkage. Overall, 42,926 hospitalized COVID-19 patients were included in the study, of which 38,715 were from Lombardy, 3,229 were from Veneto and 982 were from Reggio Emilia (figure 1). Patients’ median age was 69 years (IQR: 57–79) and 62·6% were males. In total, 11,205 (26·1%) patients died over a median follow-up of 24 days (IQR: 10–35). The median time from hospitalization to death was 6 days (IQR: 3–12); 69·4% of patients did not have any prior hospitalization reporting the comorbidities included in the Charlson Index, while 6·0% had a Charlson index ≥3 (table 1).

**Table 1.** Baseline patient characteristics, deaths and case fatality rate per 1000 person-days, by region, age, sex and Charlson index.

|  | All patients<br>N (%) | Person-time<br>(days) | N. deaths | Case fatality rate<br>(per 1,000<br>person-days) |
| --- | --- | --- | --- | --- |
| **Total** | 42,926 | 1,016,708 | 11,205 | 11·02 |
| **Catchment area** |  |  |  |  |
| Lombardy | 38,715 (90·2) | 965,129 | 10,569 | 10·95 |
| Veneto | 3,229 (7·5) | 39,640 | 439 | 11·07 |
| Reggio Emilia | 982 (2·3) | 11,939 | 197 | 16·50 |
| **Age, years** |  |  |  |  |
| 18-49 | 5,561 (13·0) | 159,252 | 141 | 0·89 |
| 50-59 | 7,172 (16·7) | 204,144 | 451 | 2·21 |
| 60-69 | 8,754 (20·4) | 233,431 | 1,484 | 6·36 |
| 70-79 | 10,953 (25·5) | 244,025 | 3,867 | 15·85 |
| 80-89 | 8,880 (20·7) | 154,177 | 4,343 | 28·17 |
| ≥90 | 1,606 (3·7) | 21,679 | 919 | 42·39 |
| **Gender** |  |  |  |  |
| Males | 26,873 (62·6) | 635,776 | 7,662 | 12·05 |
| Females | 16,053 (37·4) | 380,932 | 3,543 | 9·30 |
| **Charlson index** |  |  |  |  |
| 0 | 29,775 (69·4) | 753,537 | 5,805 | 7·70 |
| 1-2 | 10,575 (24·6) | 220,501 | 4,018 | 18·22 |
| ≥3 | 2,576 (6·0) | 42,670 | 1,382 | 32·39 |

The survival curves show that 22·0% (95% CI 21·6%–22·4%) of patients die within the second week of hospitalization and 27·6% (95% CI 27·2%–28·1%) die within 30 days. The curve then reaches a plateau with a few deaths occurring beyond one month from the start of COVID-19 hospitalization. After this point differences among regions almost disappear (tables 2 and 3). The curve had similar trajectory for all age groups, although the survival rate was higher in patients aged <50 years (2·8% died at 30 days post-hospitalization) and those aged 50-59 years (6·7% died at 30 days post-hospitalization). On the contrary, patients aged 80-89 years and ≥ 90 years had a lower survival rate (52·5% and 64·9% died at 30 days post-hospitalization, respectively). Survival rates were also higher among females both at 14- and 30-days post-hospitalization (18·7% and 23·7%, respectively). The proportion of deaths occurring at 30 days was also higher in patients with comorbidities: 20·7%, 40·2%, and 58·1% for those with a Charlson index of 0, 1–2 and ≥3, respectively (figure 2, table 3).

**Table 2.** Proportion of surviving patients at 14 days from hospitalization, estimated using the Kaplan Meier survival function, by catchment area, age, sex, and Charlson index.

|  |  | Cumulative survival rate % (95% CI) |  |  |  |  |  |
| --- | --- | --- | --- | --- | --- | --- | --- |
|  |  | Age category |  |  |  |  |  |
|  | All patients | 18-49 years | 50-59 years | 60-69 years | 70-79 years | 80-89 years | ≥90 years |
| **Overall** | 78.0 (77.6–78.4) | 98.2 (97.0–98.6) | 95.5 (95.0–96.0) | 87.1 (86–87.9) | 70.4 (69.0–71.3) | 56.8 (55.7–57.9) | 47.4 (44.9–50.0) |
| **Catchment area** |  |  |  |  |  |  |  |
| Lombardy | 77.5 (77.1–77.9) | 98.1 (97.8–98.5) | 95.4 (94.9–95.9) | 86.6 (85.9–87.4) | 69.2 (68.3–70.1) | 55.7 (54.6–56.8) | 46.3 (43.6–49.1) |
| Veneto | 85.7 (84.0–87.1) | 99.5 (98.5–100) | 97.4 (95.9–98.9) | 94.9 (92.9–96.9) | 84.2 (81.4–87.2) | 70.4 (66.5–74.5) | 56.2 (48.2–65.4) |
| Reggio Emilia | 77.3 (74.4–80.4) | .. | 93.0 (87.7–98.6) | 88.1 (83.1–93.5) | 81.0 (75.6–86.7) | 61.6 (55.4–68.4) | 47.5 (35.9–62.9) |
| **Gender** |  |  |  |  |  |  |  |
| Males | 76.1 (75.0–76.6) | 97.8 (97.0–98.3) | 94.7 (94.0–95.3) | 85.1 (84.0–86.1) | 66.9 (65.8–68.0) | 50.4 (49.0–51.8) | 42.8 (38.9–47.2) |
| Females | 81.3 (80.0–81.9) | 98.8 (98.0–99.3) | 97.1 (96.0–97.8) | 91.6 (90.5–92.7) | 77.2 (75.8–78.5) | 65.3 (63.7–66.8) | 50.2 (47.0–53.6) |
| **Charlson index** |  |  |  |  |  |  |  |
| 0 | 83.9 (83.0–84.3) | 98.6 (98.0–99.0) | 96.5 (96.0–97.0) | 90.3 (89.0–91.0) | 74.0 (72.9–75.1) | 60.2 (58.7–61.7) | 46.7 (43.2–50.6) |
| 1-2 | 67.2 (66.3–68.2) | 93.6 (91.0–96.2) | 90.2 (88.2–92.3) | 79.8 (77.9–81.7) | 66.9 (65.3–68.5) | 54.9 (53.2–56.6) | 49.9 (45.9–54.3) |
| ≥3 | 54.0 (52.0–56.0) | 80.1 (65.9–97.4) | 73.8 (64.9–83.9) | 64.4 (59.1–70.0) | 56.2 (52.8–59.8) | 49.2 (46.2–52.3) | 42.5 (35.7–50.5) |

**Abbreviation:** 95% CI = 95% confidence interval

**Table 3.**
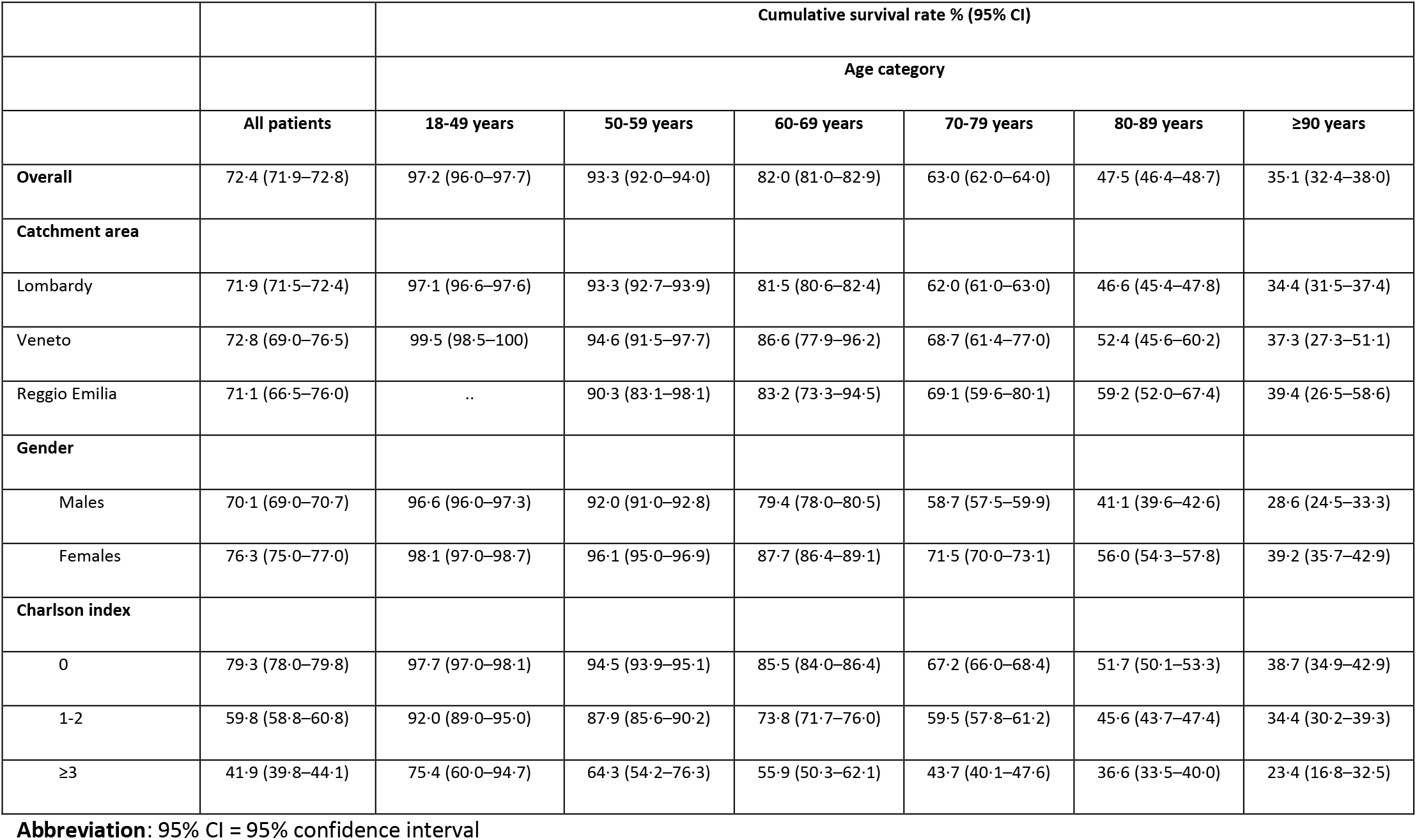
Proportion of surviving patients at 30 days from hospitalization, estimated using the Kaplan Meier survival function, by catchment area, age, sex, and Charlson index.

|  |  | Cumulative survival rate % (95% CI) |  |  |  |  |  |
| --- | --- | --- | --- | --- | --- | --- | --- |
|  |  | Age category |  |  |  |  |  |
|  | All patients | 18-49 years | 50-59 years | 60-69 years | 70-79 years | 80-89 years | ≥90 years |
| Overall | 72.4 (71.9–72.8) | 97.2 (96.0–97.7) | 93.3 (92.0–94.0) | 82.0 (81.0–82.9) | 63.0 (62.0–64.0) | 47.5 (46.4–48.7) | 35.1 (32.4–38.0) |
| Catchment area |  |  |  |  |  |  |  |
| Lombardy | 71.9 (71.5–72.4) | 97.1 (96.6–97.6) | 93.3 (92.7–93.9) | 81.5 (80.6–82.4) | 62.0 (61.0–63.0) | 46.6 (45.4–47.8) | 34.4 (31.5–37.4) |
| Veneto | 72.8 (69.0–76.5) | 99.5 (98.5–100) | 94.6 (91.5–97.7) | 86.6 (77.9–96.2) | 68.7 (61.4–77.0) | 52.4 (45.6–60.2) | 37.3 (27.3–51.1) |
| Reggio Emilia | 71.1 (66.5–76.0) | .. | 90.3 (83.1–98.1) | 83.2 (73.3–94.5) | 69.1 (59.6–80.1) | 59.2 (52.0–67.4) | 39.4 (26.5–58.6) |
| Gender |  |  |  |  |  |  |  |
| Males | 70.1 (69.0–70.7) | 96.6 (96.0–97.3) | 92.0 (91.0–92.8) | 79.4 (78.0–80.5) | 58.7 (57.5–59.9) | 41.1 (39.6–42.6) | 28.6 (24.5–33.3) |
| Females | 76.3 (75.0–77.0) | 98.1 (97.0–98.7) | 96.1 (95.0–96.9) | 87.7 (86.4–89.1) | 71.5 (70.0–73.1) | 56.0 (54.3–57.8) | 39.2 (35.7–42.9) |
| Charlson index |  |  |  |  |  |  |  |
| 0 | 79.3 (78.0–79.8) | 97.7 (97.0–98.1) | 94.5 (93.9–95.1) | 85.5 (84.0–86.4) | 67.2 (66.0–68.4) | 51.7 (50.1–53.3) | 38.7 (34.9–42.9) |
| 1-2 | 59.8 (58.8–60.8) | 92.0 (89.0–95.0) | 87.9 (85.6–90.2) | 73.8 (71.7–76.0) | 59.5 (57.8–61.2) | 45.6 (43.7–47.4) | 34.4 (30.2–39.3) |
| ≥3 | 41.9 (39.8–44.1) | 75.4 (60.0–94.7) | 64.3 (54.2–76.3) | 55.9 (50.3–62.1) | 43.7 (40.1–47.6) | 36.6 (33.5–40.0) | 23.4 (16.8–32.5) |

**Abbreviation:** 95% CI = 95% confidence interval

**Figure 2.**
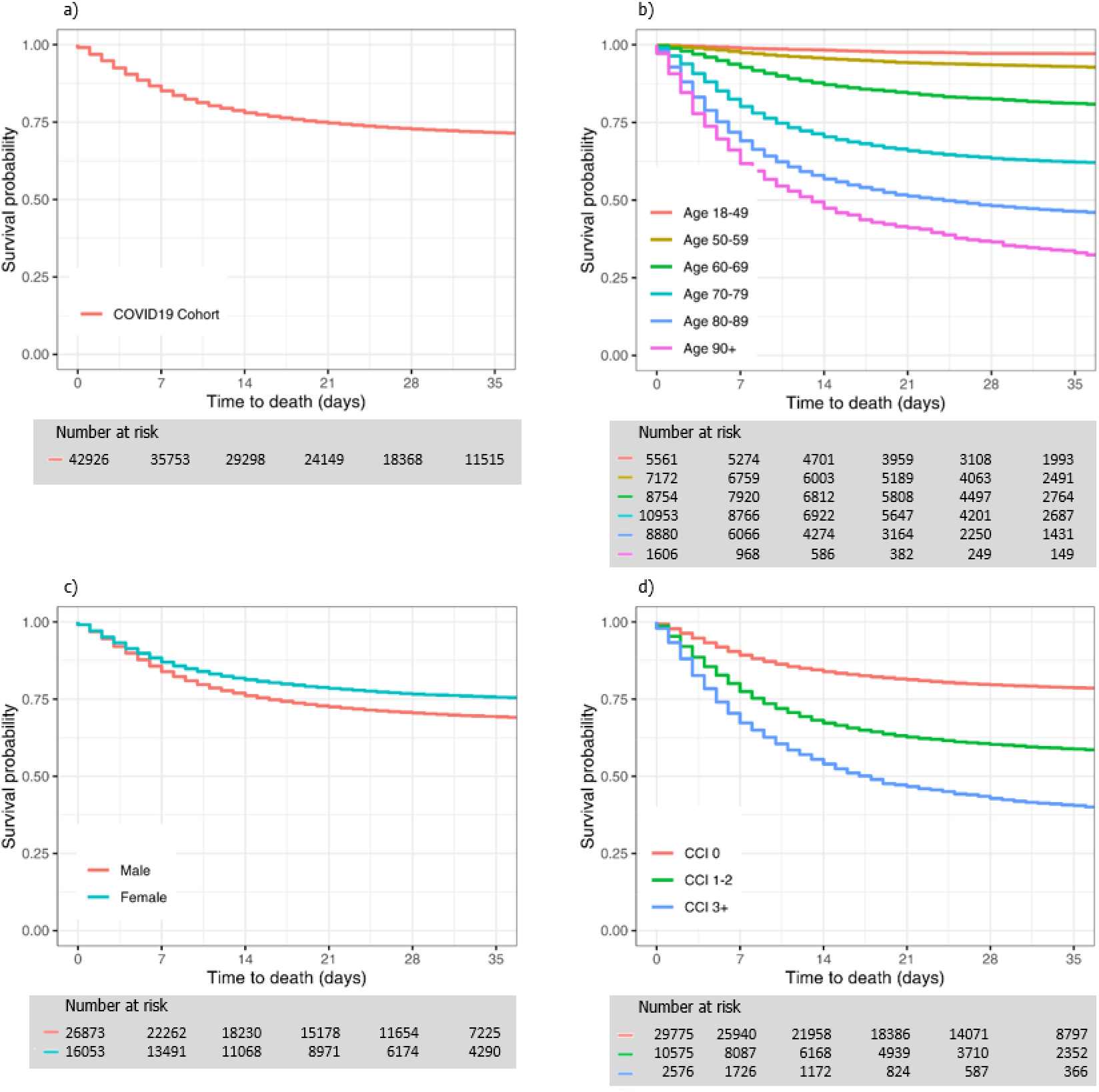
Kaplan Meier survival curves for hospitalized COVID-19 patients in Northern Italy, 21^st^ February to 21^st^ April 2020. Panel a: overall; panel b: stratified by age groups; panel c: stratified by sex at birth; panel d: stratified by Charlson index

The difference in survival between males and females was similar across all ages (tables 2 and 3, figure 3), with the HR for females vs. males ranging from 0·77 (95%CI 0·74–0·80) to 0·66 (95%CI 0·63–0·68), when adjusting for age and Charlson index (table 4). The effect of comorbidities on survival was more pronounced in younger patients, while gradually decreasing with age: in patients below the age of 50 the HR for those with Charlson index of ≥3 as compared with a Charlson index of 0 was 15·2 (95%CI 7·1–32·7), while in those over 90 it was only 1·1 (95%CI 0·93–1·4) (table 4). In younger patients (<50 years), the corresponding estimated proportion of deaths occurring at 30 days was 2·3% in those with a Charlson index of 0 and 24·6% in patients with Charlson index ≥3, while in patients over 90, these estimates were 51·3% and 76·6% in patients with a Charlson index of 0 and of ≥3, respectively (table 3, figure 4).

**Table 4.** Risk of death for hospitalized COVID-19 patients, estimated using proportional hazard Cox models for age, sex and Charlson index. Adjusted models include all reported variables and catchment area.

|  | HR (95% CI)<br>unadjusted | HR (95% CI)<br>adjusted | HR (95% CI)<br>adjusted* |
| --- | --- | --- | --- |
| **Age, years** |  |  |  |
| 18–49 | *ref* | *ref* | *ref* |
| 50–59 | 2.49 (2.06– 3.01) | 2.39 (1.98–2.89) | 2.43 (1.95– 3.02) |
| 60–69 | 7.04 (5.92– 8.37) | 6.46 (5.44–7.68) | 6.60 (5.40– 8.06) |
| 70–79 | 16.8 (14.2– 19.8) | 14.8 (12.5–17.5) | 17.8 (14.7– 21.7) |
| 80–89 | 27.6 (23.3– 32.6) | 24.3 (20.5–28.8) | 31.7 (26.1– 38.5) |
| ≥90 | 37.5 (31.4– 44.8) | 36.2 (30.3–43.3) | 52.6 (42.6– 65.0) |
| **Gender** |  |  |  |
| Males | *ref* | *ref* | *ref* |
| Females | 0.77 (0.74– 0.80) | 0.66 (0.63–0.68) | 0.65 (0.63– 0.68) |
| **Charlson index** |  |  |  |
| 0 | *ref* | *ref* | *ref* |
| 1-2 | 2.24 (2.15– 2.33) | 1.32 (1.26–1.37) | 3.74 (2.45– 5.70) |
| ≥3 | 3.65 (3.44– 3.87) | 1.76 (1.66–1.87) | 15.2 (7.08– 32.7) |
| **Gender x Charlson index** |  |  |  |
| Age 18-49 × CCI 0 | .. | .. | *ref* |
| Age 50-59 × CCI 1-2 | .. | .. | 0.67 (0.42–1.08) |
| Age 60-69 × CCI 1-2 | .. | .. | 0.53 (0.34–0.81) |
| Age 70-79 × CCI 1-2 | .. | .. | 0.34 (0.22–0.52) |
| Age 80-89 × CCI 1-2 | .. | .. | 0.31 (0.20–0.48) |
| Age ≥90 × CCI 1-2 | .. | .. | 0.26 (0.17–0.41) |
| Age 50-59 × CCI ≥3 | .. | .. | 0.46 (0.20–1.08) |
| Age 60-69 × CCI ≥3 | .. | .. | 0.25 (0.11–0.54) |
| Age 70-79 × CCI ≥3 | .. | .. | 0.13 (0.06–0.28) |
| Age 80-89 × CCI ≥3 | .. | .. | 0.09 (0.04–0.20) |
| Age ≥90 × CCI ≥3 | .. | .. | 0.07 (0.03–0.16) |

Abbreviations: CCI= Charlson index; CI= confidence interval; HR= hazard ratio; ref= reference
\* Interpretation of HR with the interaction term: in the model with the interaction terms, the HRs of CCI decline with increasing age. For instance, the HR of CCI ≥ 3 for the age class 50-59 is equal to the HR of CCI ≥ 3 (15.2) multiplied by the interaction term of CCI ≥3 and age class 50-59 (0.46) -> $15.2 \times 0.46 = 6.99$ . Similarly, HR for CCI ≥ 3 and age 70-79 is equal to $15.2 \times 0.13 = 1.98$ .

**Figure 3.**
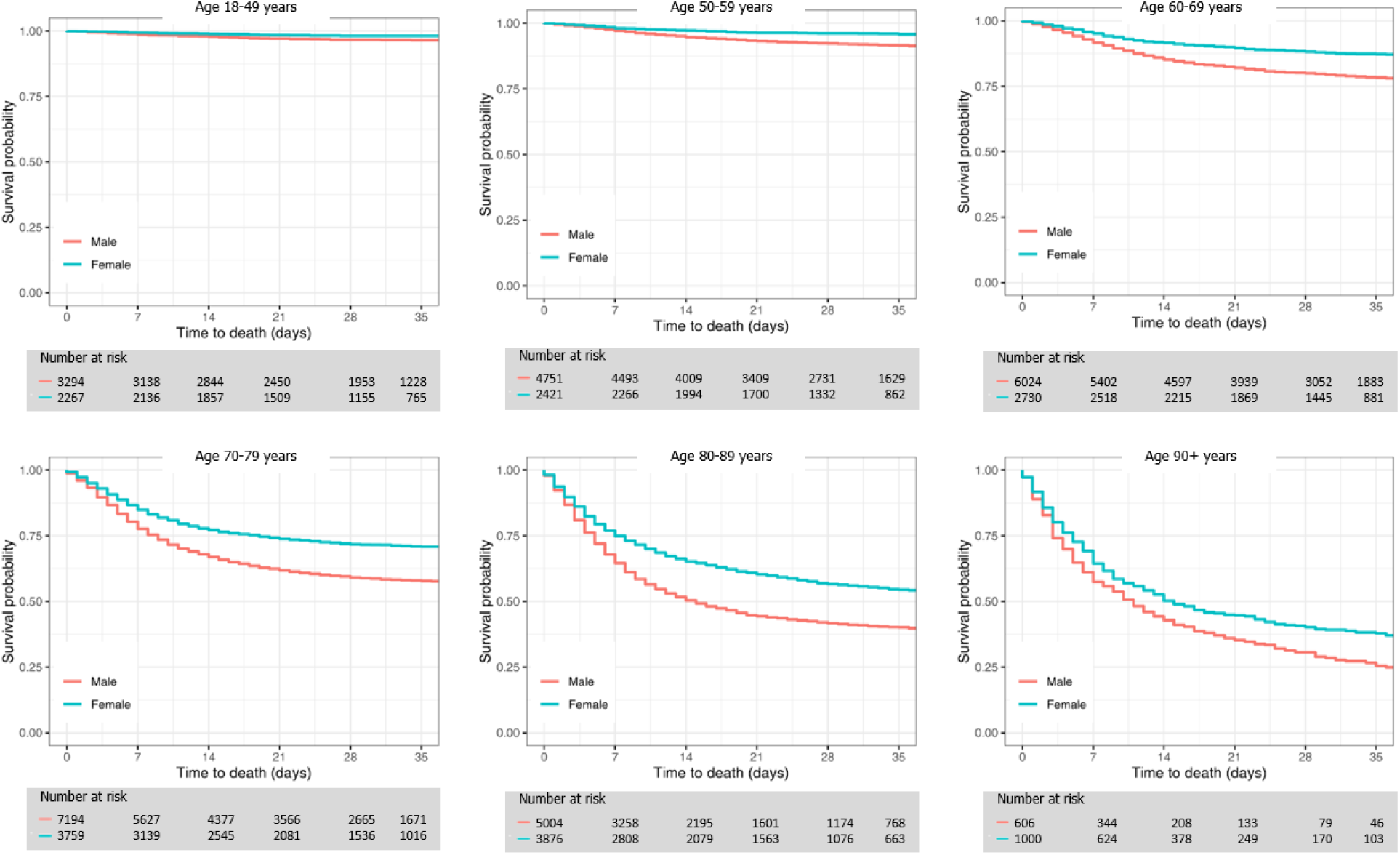
Kaplan Meier survival curves for hospitalized COVID-19 patients in Northern Italy stratified by age and sex at birth, from 21^st^ February to 21^st^ April 2020

## Discussion

In a large population-based cohort of hospitalized COVID-19 patients we observed a 27·6% fatality rate at one month after hospitalization, which reached a plateau thereafter. The fatality rate was slightly higher in males (29·9%) than in females (25·7%), increasing dramatically to 52% in persons aged 80-90 and 65% in persons aged over 90. The burden of comorbidities, measured using the Charlson index, is an important prognostic factor in younger patients, but after the age of 80 the impact of this on fatality rate is smaller, as probably other factors, such as frailty, become more important risk factors for death.

Compared to previous studies and current statistics, we observed a much higher CFR in hospitalized cohorts, where deaths rates rarely reach 20%.^13,14,17^ A recent study on 16,749 UK patients hospitalized with COVID-19 found a CFR of 33%.^22^ The differences between the UK study and the present study could be explained by the different criteria used for deciding which patients should be hospitalized, but our data are in line with the very high fatality rate observed in all settings in Italy, not only in hospitalized patients.^9,15^ Several hypotheses have been proposed to explain differences in CFR in various countries, including different viral strains that could cause more or less severe disease, the different capacity of a healthcare system to respond to the COVID-19 crisis, as well as different case definitions, with some countries including probable cases in the overall death toll. The CFR can increase in some areas if there is a surge of infected patients, which adds to the strain on the healthcare system and can overwhelm its medical resources.^23^ Data from hospitalized COVID-19 patients in New York City show a CFR of 20% at approximately 6 days of follow-up;^19^ although this figure seems much higher than our observed survival in the first week after hospital admission, if we consider that the average Charlson index in the New York population was over 4, the fatality rate is consistent with our estimate for the population with a Charlson index ≥3, reported in figure 2, which ranges between 20 and 25%. An relatively long follow-up and the very old age of the hospitalized population may partly explain the high fatality observed in the Italian cohort.

Our results confirm a lower survival in males, even after adjusting for comorbidities.^24^ Interestingly, males had a higher probability of having COVID-19 as well as of being hospitalized^15,25^ and dying after hospitalization due to COVID-19, according to our data. Therefore, the excess morbidity and mortality is underestimated when we observe only hospitalized patients. Different hypotheses involving the role of ACE2 as principal receptor of SARS-CoV-2 for its entry into cells have been proposed to explain the higher susceptibility to infection and the lower survival in males.^26^ The role of the TMPRSS2, a protease involved in virus binding and up-regulated by androgens, has been also proposed.^27^

The impact of age on the risk of death after developing COVID-19 was dramatic in our cohort, in line with all previous studies.^7,15^ Again, the impact of age on survival could be partially underestimated when studying hospitalized cohorts, as older patients, at least until the age of 80, also have a higher risk of being hospitalized. The excess risk cannot be entirely attributed to the underlying comorbidities that are usually more frequent in older patients, as can be deduced comparing the survival curves of people with the same Charlson index in different age groups (figure 4). On the other hand, severe symptoms in children and adolescents seem extremely rare, and very few cases of death have been reported for children aged less than 10 years so far.^8,28^

**Figure 4.**
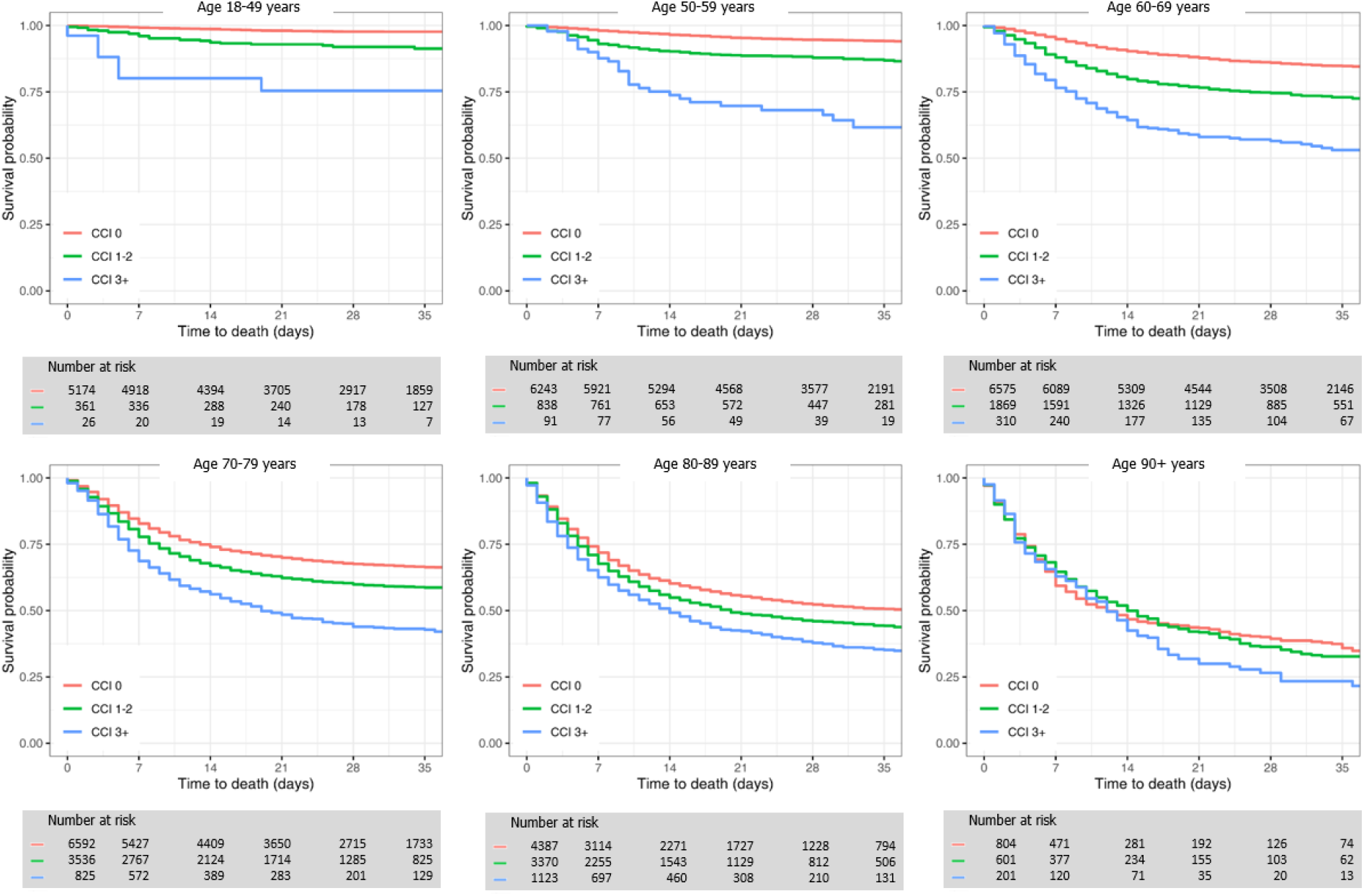
Kaplan Meier survival curves for hospitalized COVID-19 patients in Northern Italy stratified by age and Charlson index, from 21^st^ February to 21^st^ April 2020

The burden of comorbidities play an important role in determining the risk of death in COVID-19 patients in our cohort, confirming the results of all previous studies.^17,18^ The large number of cases identified allowed us to study the effect of comorbidity burden across age groups, giving a precise survival estimate by comorbidity in each age group. The effect of comorbidities on survival is much more pronounced in younger patients than in older ones. In our study design, the assessment of the pre-existing comorbidities is independent from the outcome being collected through the analysis of patient use of healthcare services in the years preceding the epidemic. Therefore, even if we may underestimate the presence of comorbidities, this misclassification is not differential in severe or fatal cases and in non-severe cases, thus the hazard ratios should be unbiased.

The availability of life-sustaining therapies or lack thereof should be considered when interpreting the COVID-19 mortality rate: poor outcomes may be due to known risk factors such as old age, frailty, comorbidities, profound disability, or because of effects of logistic limitations associated with lack of medical staff and medical staff burn-out, lack of beds and/or medical supplies. Moreover, although patients might indeed have SARS-CoV-2 infection, the infection itself may not necessarily be the cause of death. Extreme examples include patients with metastatic cancer or terminal organ failure.^12^

Due to the regional structure of the Italian NHS, the responses to the COVID-19 epidemic vary from region to region. The Lombardy region is home to a sixth of the Italian population (10·08 million inhabitants) and accounts for 37% of cases and 53% of deaths of the country, as of 21^st^ April, 2020. The COVID-19 outbreak initially hit Italy in two small towns, Codogno and Vò Euganeo, in Lombardy and Veneto, respectively. The two outbreaks developed differently: in Lombardy the spread of infection was quick and several other clusters emerged rapidly;^4^ in Veneto, the first cluster had a limited spread until the diffusion of cases in all Northern Italian regions occurred. Finally, in Reggio Emilia the outbreak was related to the expansion of the first cluster occurring in the neighboring area of Codogno. Differences in the pattern of spread of the infection across the three areas are probably due to the different size of the initial clusters when they emerged, i.e. much larger in Codogno than in Vò Euganeo, but also on the epidemic control strategies which were implemented initially: Veneto opted for strict containment of the outbreak and piloted mass testing in selected areas, whereas Lombardy expanded hospital services to meet a massively increased need for hospitalization and beds in ICUs.^29^ As result the proportion of positive tests on the total number of swabs performed was 20% in Lombardy, 5·1% in Veneto and 13·9% in Emilia-Romagna. Clinical criteria which must be met for COVID-19 hospital admission may vary among regions, principally due to a different organization of primary care and the availability of hospital beds, which varied in different places and moments of the epidemic, and availability of other options for managing less severe patients, such as home-based treatment with monitoring. Where special units for home-based care for COVID-19 patients were activated, patients were treated and followed-up at home, when possible. Nevertheless, differences in the overall CFRs among the three catchment areas at 30 days post-hospitalization in our cohort were modest, suggesting the generalizability of the results once the plateau is reached.

### Implications for practice

The aim of this study was to provide a precise estimate of survival over time after hospitalization for COVID-19 patients. Survival and CFR are very useful also for the design of phase II clinical trials and to calculate the adequate sample size for phase III clinical trials. We found a very high CFR, particularly in older people. Since this endpoint is very common, we suggest that future experimental studies investigating potential therapeutic interventions in COVID-19 patients should prioritize survival as a main outcome. Furthermore, we found that the fatality rate reaches a plateau 30 days after hospitalization, thus implying that studies should have at least one month of follow-up after COVID-19 hospitalization to properly observe deaths; studies with shorter follow-up may not capture all deaths related to COVID-19, ultimately leading to the overestimation of the potential benefit of COVID-19 interventions in phase II studies in absence of a comparator.

Hard outcomes such as the CFR have a crucial role and, together with survival estimates, should guide health-care leaders and policy makers in developing public health strategies at national and international levels.^23^

## Acknowledgments

No funding was received for the conduct of the study.

## Authors and contributors

PGR, EF, MMas, GT and SSA contributed to study design, data collection, data analysis, data interpretation, the literature search, and first draft writing of the manuscript.

JS edited the final version of the manuscript, contributed to literature search and data visualization.

GP, OL, DC contributed to study design data, data collection, interpretation and, the literature search.

MMas, MMar, MP contributed to data collection, software programming, data analysis and data interpretation.

All authors reviewed and approved the final version of the report.

The corresponding author had full access to all the data in the study and had final responsibility for the decision to submit for publication.

